## Supplementary Figures for "Generalized Radiograph Representation Learning via Cross-supervision between Images and Free-text Radiology Reports"

### Supplementary information

#### Figures

### Comparison with self-supervised and transfer learning baselines

#### a. NIH ChestX-ray dataset

| Training data | Method | Mean | Atelectasis | Cardiomegaly | Consolidation | Edema | Effusion | Emphysema | Fibrosis | Hernia | Infiltration | Mass | Nodule | Pleural Thickening | Pneumonia | Pneumothorax | p-value |
|---|---|---|---|---|---|---|---|---|---|---|---|---|---|---|---|---|---|
| 0.8k (1%) | Our REFERS | **76.7** | **77.5** | **85.6** | **78.6** | **84.9** | **85.4** | **79.5** | **72.3** | **77.1** | **67.5** | **76.2** | **66.5** | **71.6** | **69.3** | **81.7** | 8.35e-4 |
| | Model Genesis | 70.3 | 72.1 | 67.1 | 75.8 | 76.1 | 80.6 | 72.6 | 64.8 | 73.5 | 65.7 | 65.2 | 62.2 | 67.6 | 64.8 | 76.2 | |
| | C2L | 71.0 | 75.1 | 67.1 | 77.6 | 75.1 | 83.4 | 71.5 | 66.8 | 70.0 | 63.8 | 70.1 | 66.2 | 68.1 | 65.7 | 74.4 | |
| | Context Restoration | 67.8 | 69.1 | 64.4 | 73.2 | 73.8 | 78.1 | 70.0 | 62.1 | 70.2 | 65.2 | 62.4 | 59.1 | 65.0 | 62.2 | 73.8 | |
| | TransVW | 71.2 | 74.5 | 68.9 | 76.7 | 79.8 | 81.1 | 67.9 | 68.7 | 68.2 | 66.8 | 66.5 | 66.2 | 68.5 | 68.8 | 75.0 | |
| | ImageNet Pre-training | 69.8 | 73.3 | 69.6 | 76.0 | 81.7 | 80.5 | 67.1 | 64.9 | 64.8 | 65.8 | 67.0 | 62.3 | 65.7 | 65.0 | 74.0 | |
| 8k (10%) | Our REFERS | **80.9** | **80.1** | **89.8** | **79.5** | **87.8** | **87.5** | **88.2** | **77.2** | **86.1** | **69.6** | **82.0** | **72.8** | **74.2** | **72.2** | **85.6** | 8.72e-4 |
| | Model Genesis | 75.7 | 77.2 | 72.8 | 77.5 | 85.7 | 85.2 | 81.0 | 75.3 | 78.0 | 68.4 | 73.1 | 69.5 | 72.2 | 67.7 | 80.4 | |
| | C2L | 76.6 | 78.0 | 75.5 | 77.5 | 84.1 | 85.7 | 81.2 | 73.7 | 79.5 | 67.4 | 77.5 | 71.7 | 72.0 | 67.3 | 81.9 | |
| | Context Restoration | 73.9 | 75.5 | 70.6 | 77.1 | 84.5 | 84.2 | 79.4 | 73.1 | 67.5 | 68.1 | 70.9 | 66.9 | 71.7 | 65.2 | 79.1 | |
| | TransVW | 74.3 | 76.5 | 70.8 | 77.6 | 83.0 | 84.8 | 79.7 | 69.9 | 74.7 | 68.5 | 72.1 | 68.3 | 72.4 | 63.2 | 79.6 | |
| | ImageNet Pre-training | 74.4 | 74.2 | 79.8 | 75.9 | 85.7 | 83.2 | 80.4 | 72.1 | 74.0 | 64.1 | 71.7 | 65.6 | 69.6 | 66.2 | 79.7 | |
| 80k (100%) | Our REFERS | **84.7** | **83.0** | **92.3** | **82.1** | **90.2** | **88.7** | **91.4** | **83.9** | **93.3** | **74.1** | **85.5** | **76.7** | **78.5** | **77.0** | **89.1** | 1.94e-3 |
| | Model Genesis | 81.0 | 78.8 | 84.5 | 79.2 | 87.8 | 86.6 | 89.7 | 81.0 | 85.2 | 71.1 | 81.9 | 73.2 | 75.8 | 73.0 | 85.6 | |
| | C2L | 82.2 | 81.1 | 90.2 | 81.0 | 88.1 | 88.0 | 88.3 | 80.8 | 86.8 | 72.0 | 82.7 | 74.1 | 76.2 | 75.3 | 85.9 | |
| | Context Restoration | 78.7 | 75.8 | 82.9 | 76.4 | 86.6 | 84.8 | 88.2 | 78.6 | 83.0 | 70.0 | 79.6 | 69.5 | 73.2 | 69.4 | 84.0 | |
| | TransVW | 81.7 | 79.8 | 85.0 | 80.0 | 88.2 | 87.1 | 90.1 | 81.8 | 85.9 | 72.3 | 82.6 | 74.4 | 76.6 | 74.0 | 86.1 | |
| | ImageNet Pre-training | 80.0 | 78.3 | 89.3 | 77.6 | 87.9 | 85.9 | 87.4 | 78.5 | 88.8 | 65.9 | 79.9 | 70.7 | 74.5 | 71.0 | 84.7 | |

#### b. VinBigData Chest X-ray Abnormalities Detection

| Training data | Method | Mean | Aortic Enlargement | Atelectasis | Calcification | Cardiomegaly | Consolidation | ILD | Infiltration | Lung Opacity | Nodule-Mass | Other Lesion | Pleural Effusion | Pleural Thickening | Pneumothorax | Pulmonary Fibrosis | p-value |
|---|---|---|---|---|---|---|---|---|---|---|---|---|---|---|---|---|---|
| 0.1k (1%) | Our REFERS | **83.0** | 88.4 | **85.1** | **71.2** | **91.4** | **88.6** | **83.7** | **81.9** | **86.3** | **74.7** | **77.7** | **86.0** | **81.8** | **85.7** | **79.4** | 8.72e-5 |
| | Model Genesis | 70.7 | 87.8 | 65.7 | 57.6 | 87.5 | 72.9 | 67.0 | 64.3 | 63.6 | 67.7 | 68.7 | 77.5 | 77.7 | 63.1 | 68.4 | |
| | C2L | 75.3 | **89.1** | 71.3 | 64.5 | 88.7 | 77.4 | 71.2 | 70.3 | 70.2 | 72.5 | 75.0 | 81.4 | 80.7 | 69.5 | 72.5 | |
| | Context Restoration | 67.9 | 74.4 | 63.6 | 63.7 | 74.7 | 71.2 | 68.6 | 67.5 | 70.0 | 64.2 | 65.4 | 67.7 | 68.0 | 64.8 | 66.5 | |
| | TransVW | 73.6 | 82.8 | 68.4 | 68.5 | 83.4 | 74.7 | 73.6 | 72.6 | 75.8 | 69.0 | 70.3 | 74.1 | 75.1 | 68.7 | 73.5 | |
| | ImageNet Pre-training | 69.7 | 77.7 | 64.5 | 67.3 | 80.0 | 70.1 | 69.6 | 68.1 | 73.3 | 64.9 | 67.2 | 69.3 | 70.3 | 62.7 | 70.1 | |
| 1k (10%) | Our REFERS | **88.2** | **92.6** | **89.6** | 78.4 | **92.9** | **94.4** | **86.7** | **87.4** | **91.2** | **83.6** | **84.7** | **90.2** | **88.1** | **89.6** | **85.8** | 4.34e-4 |
| | Model Genesis | 82.7 | 88.6 | 83.3 | 78.2 | 86.6 | 87.4 | 79.4 | 81.9 | 83.6 | 79.9 | 82.3 | 83.5 | 85.0 | 74.9 | 83.2 | |
| | C2L | 83.3 | 92.1 | 80.3 | **78.6** | 89.5 | 82.6 | 81.8 | 82.6 | 84.7 | 80.9 | 83.5 | 85.0 | **88.1** | 72.1 | 85.0 | |
| | Context Restoration | 82.4 | 91.4 | 75.1 | 72.4 | 89.6 | 81.6 | 80.7 | 79.6 | 85.5 | 80.7 | 82.6 | 86.0 | 87.6 | 76.6 | 84.7 | |
| | TransVW | 83.8 | 91.7 | 80.3 | 72.5 | 89.7 | 85.9 | 81.3 | 82.0 | 85.7 | 82.0 | 84.4 | 86.7 | 87.6 | 76.9 | 85.0 | |
| | ImageNet Pre-training | 82.9 | 91.6 | 81.1 | 73.3 | 89.6 | 87.8 | 79.6 | 82.1 | 85.7 | 82.4 | 84.0 | 86.6 | 87.5 | 75.1 | 85.5 | |
| 10k (100%) | Our REFERS | **90.1** | **93.6** | **90.2** | 80.4 | **93.6** | **95.1** | **91.2** | **90.3** | **93.2** | **84.6** | **86.9** | **92.3** | **90.5** | **89.7** | **89.5** | 9.33e-4 |
| | Model Genesis | 85.8 | 90.8 | 84.8 | 79.0 | 89.4 | 89.0 | 81.6 | 82.6 | 85.6 | 81.4 | 85.0 | 86.3 | 86.5 | 80.3 | 84.4 | |
| | C2L | 85.9 | 92.5 | 82.5 | **80.9** | 90.6 | 86.3 | 85.0 | 85.2 | 87.1 | 82.1 | 84.7 | 87.2 | 88.5 | 82.6 | 86.9 | |
| | Context Restoration | 83.8 | 92.7 | 76.1 | 73.0 | 90.9 | 84.2 | 81.7 | 81.3 | 86.8 | 82.0 | 84.6 | 87.1 | 87.9 | 80.8 | 84.8 | |
| | TransVW | 86.2 | 92.0 | 81.7 | 76.0 | 90.1 | 86.2 | 86.6 | 88.0 | 87.1 | 83.5 | 85.3 | 88.3 | 88.8 | 86.0 | 86.6 | |
| | ImageNet Pre-training | 84.5 | 92.6 | 81.4 | 75.9 | 91.5 | 88.3 | 80.5 | 83.0 | 86.6 | 82.7 | 84.2 | 87.2 | 87.7 | 79.9 | 86.0 | |

Figure 1: Comparison with self-supervised learning and transfer learning baselines. Note that for the sake of fairness, all baselines use the same transformer-based backbone as the radiograph transformer of REFERS (i.e., a ViT-like architecture plus the recurrent concatenation operator). Each p-value is calculated between our REFERS and the best performing baseline. The evaluation metric is Area under the ROC Curve (AUC). Best results are bolded.

### Comparison with label-supervised learning

#### a. NIH ChestX-ray dataset

| Training data | Method | Mean | Atelectasis | Cardiomegaly | Consolidation | Edema | Effusion | Emphysema | Fibrosis | Hernia | Infiltration | Mass | Nodule | Pleural Thickening | Pneumonia | Pneumothorax | p-value |
|---|---|---|---|---|---|---|---|---|---|---|---|---|---|---|---|---|---|
| 0.8k (1%) | LSP (Transformer) | 74.2 | 75.5 | 85.0 | 77.2 | **85.0** | 85.3 | 71.5 | 70.6 | 64.5 | 66.8 | 72.4 | 66.0 | 69.4 | 68.8 | 80.4 | 3.25e-3 |
| | LSP (ConvNet) | 65.8 | 66.9 | 62.4 | 71.3 | 72.1 | 76.2 | 68.0 | 60.1 | 67.4 | 64.6 | 60.3 | 56.8 | 63.1 | 60.1 | 71.9 | |
| | Our REFERS | **76.7** | **77.5** | **85.6** | **78.6** | 84.9 | **85.4** | **79.5** | **72.3** | **77.1** | **67.5** | **76.2** | **66.5** | **71.6** | **69.3** | **81.7** | |
| 8k (10%) | LSP (Transformer) | 78.2 | 77.9 | 86.3 | 77.7 | 87.2 | 85.5 | 83.8 | 76.0 | 80.2 | 67.3 | 76.0 | 69.7 | 73.0 | 71.4 | 82.4 | 2.89e-3 |
| | LSP (ConvNet) | 74.5 | 76.2 | 71.4 | 77.0 | 85.0 | 84.6 | 80.0 | 74.0 | 69.5 | 68.0 | 71.7 | 67.9 | 72.2 | 66.1 | 79.6 | |
| | Our REFERS | **80.9** | **80.1** | **89.8** | **79.5** | **87.8** | **87.5** | **88.2** | **77.2** | **86.1** | **69.6** | **82.0** | **72.8** | **74.2** | **72.2** | **85.6** | |
| 80k (100%) | LSP (Transformer) | 82.1 | 80.1 | 90.0 | 80.2 | 89.2 | 87.3 | 88.7 | 81.1 | 89.4 | 70.0 | 81.3 | 74.5 | 76.8 | 75.5 | 85.4 | 5.23e-3 |
| | LSP (ConvNet) | 81.9 | 80.2 | 85.3 | 80.5 | 88.4 | 87.4 | 90.3 | 82.1 | 86.2 | 70.0 | 83.0 | 74.9 | 77.0 | 74.6 | 86.3 | |
| | Our REFERS | **84.7** | **83.0** | **92.3** | **82.1** | **90.2** | **88.7** | **91.4** | **83.9** | **93.3** | **74.1** | **85.5** | **76.7** | **78.5** | **77.0** | **89.1** | |

#### b. VinBigData Chest X-ray Abnormalities Detection

| Training data | Method | Mean | Aortic Enlargement | Atelectasis | Calcification | Cardiomegaly | Consolidation | ILD | Infiltration | Lung Opacity | Nodule-Mass | Other Lesion | Pleural Effusion | Pleural Thickening | Pneumothorax | Pulmonary Fibrosis | p-value |
|---|---|---|---|---|---|---|---|---|---|---|---|---|---|---|---|---|---|
| 0.1k (1%) | LSP (Transformer) | 78.5 | 86.1 | 73.3 | 62.2 | 90.4 | 87.3 | 82.7 | 80.8 | 83.4 | 68.5 | 72.8 | 84.0 | 76.5 | 76.4 | 74.6 | 3.56e-4 |
| | LSP (ConvNet) | 76.0 | **89.2** | 71.1 | 64.2 | 88.9 | 78.4 | 72.4 | 70.7 | 70.9 | 72.8 | 75.4 | 81.8 | 80.9 | 74.4 | 72.8 | |
| | Our REFERS | **83.0** | 88.4 | **85.1** | **71.2** | **91.4** | **88.6** | **83.7** | **81.9** | **86.3** | **74.7** | **77.7** | **86.0** | **81.8** | **85.7** | **79.4** | |
| 1k (10%) | LSP (Transformer) | 85.8 | 89.8 | 84.5 | 75.6 | 91.9 | 91.9 | 86.4 | **87.8** | 88.2 | 81.0 | 82.0 | 87.6 | 84.8 | 86.9 | 84.5 | 8.69e-4 |
| | LSP (ConvNet) | 85.2 | 91.7 | 86.1 | 76.9 | 89.7 | 84.8 | 85.4 | 83.6 | 85.8 | 82.6 | 84.5 | 86.1 | 88.5 | 81.3 | **85.8** | |
| | Our REFERS | **88.1** | **92.6** | **89.6** | **78.4** | **92.9** | **94.4** | **86.7** | 87.4 | **91.2** | **83.6** | **84.7** | **90.2** | **88.1** | **89.6** | **85.8** | |
| 10k (100%) | LSP (Transformer) | 87.6 | 92.4 | 86.5 | 76.1 | 92.8 | 92.3 | 90.4 | 88.4 | 89.1 | 83.1 | 84.0 | 89.2 | 87.6 | 88.6 | 86.0 | 1.05e-3 |
| | LSP (ConvNet) | 87.2 | 92.8 | 87.7 | 79.3 | 91.3 | 88.7 | 87.9 | 86.0 | 88.1 | 83.2 | 85.7 | 88.1 | 88.7 | 86.0 | 86.8 | |
| | Our REFERS | **90.1** | **93.6** | **90.2** | **80.4** | **93.6** | **95.1** | **91.2** | **90.3** | **93.2** | **84.6** | **86.9** | **92.3** | **90.5** | **89.7** | **89.5** | |

Figure 2: Comparison with methods using human-assisted structured labels. Note that for fairness, both LSP (Transformer) and REFERS share the same transformer-based backbone (i.e., the ViT architecture plus the recurrent concatenation operator). Each p-value is calculated between the results from our REFERS and LSP (Transformer). The evaluation metric is Area under the ROC Curve (AUC). Best results are bolded.
